# Bicuspid Aortic Valve in Heritable Thoracic Aortic Disease: Insights from the Montalcino Aortic Consortium

**DOI:** 10.1101/2025.11.26.25341124

**Authors:** Kishan L. Asokan, Neha Muraly, Dongchuan Guo, Deep M. Shah, Ernesto C. Martinez, Elena Cervi, Julie De Backer, Laura Muiño Mosquera, Wannes Renders, Alan C. Braverman, Michelle Lim, Maral Ouzonian, Richmond W. Jeremy, Shaine A. Morris, Irina V. Volguina, Talha Niaz, Anji T. Yetman, David W. Jantzen, Gisela Teixido-Tura, Arturo Evangelista, Guillaume Jondeau, Olivier Milleron, Dianna M. Milewicz, Siddharth K. Prakash

## Abstract

Bicuspid aortic valve (BAV) is the most common congenital heart malformation and predisposes to thoracic aortic aneurysms. The Montalcino Aortic Consortium (MAC) registry collects genetic and phenotypic data on individuals with heritable thoracic aortic disease genes (HTAD PVs) that primarily regulate smooth muscle cell contraction or TGF-β signaling (TGF-β PVs, termed Loeys-Dietz Syndrome (LDS)). We evaluated associations between BAV, HTAD PVs and aortic outcomes in the MAC cohort.

BAV was present in 48 (6%) of 816 MAC participants (age 38 [IQR 20-52] years, 51% female) and was not significantly increased in males or in the entire TGF-β PV group (417 with TGF-β PVs, p=0.14) but was enriched in participants with *TGFBR1* or *TGFBR2* PVs (16/168, 10% vs. 32/648, 5%, PR 1.93 [1.08, 3.43], p<0.05). BAV was associated with aortic regurgitation (AR, 17/46, 37% vs. 84/752, 11%, PR 3.3 [2.2-5.1], *P<*0.001), younger age at first aortic event (24 vs. 40 years, p<0.001), primarily reflecting increased proximal aortic repair (27/48, 56% vs. 227/768, 30%, PR 1.90 [1.45, 2.50], *P<*0.0001), but not aortic dissection (6/48, 13% vs. 134/768, 18%, P>0.5). Sinus of Valsalva dilation (Z>3, 25/48, 52% vs. 128/768, 17%, *P<*0.0001) and ascending aorta dilation (Z>3, 12/48, 25% vs. 26/768, 3%, *P<*0.0001) were more common in participants with BAV. Participants with both TGF-β PV and BAV more frequently had aortic valve surgery (17/30, 57% vs. 110/387, 28%, PR 1.99 [1.40-2.83], p<0.01) and sinus of Valsalva dilation (20/29, 69% vs. 91/258, 35% PR 1.96 [1.40-2.63], p<0.001), but not ascending dilation, than participants with TGF-β PV who did not have BAV.

BAV is enriched in individuals with HTAD PV, most prominently in the subgroup with *LDS TGFBR1* and *TGFBR2*, and is associated with accelerated aortic valve and aortic diseasebut does not confer an increased risk for aortic dissection. These observations highlight the potential benefits of genetic testing for BAV patients who have a family history of HTAD, a sinus of Valsalva aneurysm, or clinical features suggestive of LDS.

## Introduction

Bicuspid aortic valve (BAV) is the most common adult congenital heart malformation (0.7-1.3% in Europeans) and is three times more common in men than in women [1, 2]. BAV is characterized by variable valvular phenotypes, aortic manifestations, and associated complications [3, 4]. In many individuals, BAV is never diagnosed during life and does not cause clinical complications [5]. The most common clinical presentation of BAV is incidentally discovered and slowly progressive aortic valve disease with or without thoracic aortic dilation. Affected patients often require longitudinal surveillance because more than half eventually require aortic valve or aortic interventions [6–8]. In less than 1% of cases, BAV is associated with a genetic syndrome, most commonly Turner syndrome or Loeys-Dietz syndrome [9].

BAV is phenotypically diverse. The typical BAV configuration (90-95% of cases) consists of two aortic cusps separated by a single fibrocalcific raphe due to incomplete separation of the right and left coronary cusps (70-80%), the right and non-coronary cusps (20-30%), or the left and non-coronary cusps (3-6%) [4,10]. In 5-7% of cases there are 2 symmetrical cusps without a raphe. Aortic valve manifestations and complications such as aortic valve stenosis (AS), regurgitation (AR), and endocarditis can vary depending on valve configuration [11]. Aortic dilation is present in more than 70% of individuals with BAV. The overall risk for dissection in BAV cohorts appears to be very low (0.1%) but is significantly higher than the general population [6,12].

Heritable thoracic aortic disease (HTAD) refers to thoracic aortic aneurysms and aortic dissections that are primarily caused by dominantly inherited genetic variants [13]. HTAD genes encode proteins regulating TGF-β signaling (*TGFBR1*, *TGFBR2*, *TGFB2*, *TGFB3, SMAD3, SMAD2*), the formation and stability of the extracellular matrix (*FBN1*, *MFAP5*, *LOX*, *COL3A1*), and smooth muscle cell contraction (*ACTA2*, *MYLK*, *PRKG1*, *MYH11*). Although HTAD may be associated with genetic syndromes, most patients have no syndromic features [14]. The risk of aortic complications and the age of onset vary according to the mutated HTAD gene. Early diagnosis, close surveillance, and preventative intervention can significantly improve life expectancy [15–16]. The Montalcino Aortic Consortium (MAC) was founded in 2013 as an international collaboration of translational investigators with the primary goal of understanding the natural history of individuals with pathogenic variants (PVs) in HTAD genes. The MAC registry data was instrumental to define the clinical features and dissection risk associated with different HTAD genes which informed the 2022 clinical guideline for HTAD management [17–19].

Although BAV may be inherited in an autosomal dominant pattern, the genetic architecture is complex and involves multiple interacting genes [20]. HTAD PVs are rarely identified in patients with BAV, even with complex valvulo-aortopathy phenotypes. These observations are consistent with studies that found minimal genetic overlap between HTAD and BAV [21]. However, there is evidence that TGF-β signaling can modify BAV disease by regulating aortic valve differentiation and left ventricular outflow tract development [22]. We hypothesize that BAV may interact with HTAD genotypes and sex to modify aortic and valvular disease, causing more pronounced aortic and valvular phenotypes. This interaction may accelerate the progression of aortic disease with increased risks for aortic dissection or aortic surgery. The aims of this study were to describe the prevalence of BAV in MAC registry participants by genotype and sex and to investigate the prognostic impact of BAV on associated clinical features and adverse cardiovascular outcomes.

## Methods

We conducted a multicenter study of participants in the Montalcino Aortic Consortium (MAC) registry. The study protocol was approved by the Committee for the Protection of Human Subjects at the University of Texas Health Science Center at Houston (HSC-MS-16-0191). Informed consent was obtained from all participants or obtained with a waiver of consent.

De-identified clinical data was extracted from a deidentified registry database [23]. All variants were interpreted according to American College of Medical Genetics and Genomics standards and guidelines supplemented by relevant gene- and disease-specific criteria [24]. MAC participants with pathogenic and likely pathogenic variants were combined into one group (PV) for analysis. MAC allows for inclusion of participants with variants of uncertain significance (VUSs) if there is clinical suspicion for contribution to HTAD based on phenotype and segregation. We selected a subset of VUSs for analysis with the PV group based on an independent review considering genetic evidence, population frequency, computational predictions, and available functional data. These interpretations reflect research-based assessments made in the context of this study and are not intended as formal clinical reclassifications. Individuals without a validated HTAD gene variant or complete clinical data were excluded from the analysis.

The primary covariates were sex, HTAD PV, aortic stenosis (AS); mild, moderate, or severe aortic regurgitation (AR); mild, moderate, or severe mitral regurgitation (MR), or mitral valve prolapse (MVP). The primary outcome variables were any aortic dilation, aortic dissection, aortic or aortic valve surgery, or thoracic aortic aneurysm, defined as a sinus or ascending Z-score > 3. Secondary outcomes included symptomatic arrhythmias or a clinical diagnosis of congestive heart failure (CHF).

Valve morphology was verified by direct review of echocardiographic images and was classified according to international consensus criteria in parasternal short-axis views [4]. Experienced cardiologists at contributing MAC sites scored images according to American Society of Echocardiography guidelines [25].

Categorical data were compared using chi-squared or Fisher’s exact tests and reported as prevalence ratios (PR). Continuous data was compared using unpaired t-tests when data were normally distributed. We did not conduct a survival analysis because most MAC participants (81%) experienced their first aortic event prior to enrollment.

## Results

The MAC registry database included 1485 participants at the time of this study. After 242 participants were excluded due to incomplete genetic data and 270 participants were excluded due to incomplete phenotypic data, a total of 973 participants were included in the primary analysis. Based on a structured review of available evidence, 84 VUSs were reclassified as likely pathogenic or pathogenic under the ACMG framework (Table S1). The study cohort consisted of 816 MAC participants with PVs (417 with TGF-β PVs, 208 with ECM PVs, 171 with SMC PVs, 48 with BAV, 38 [IQR 20-52] years, 51% female, 53% probands) and 157 participants with VUSs (16 with BAV). Images of acceptable quality were available from 33 participants with BAV to assess aortic valve morphology (Figure 1).

**Figure 1.**
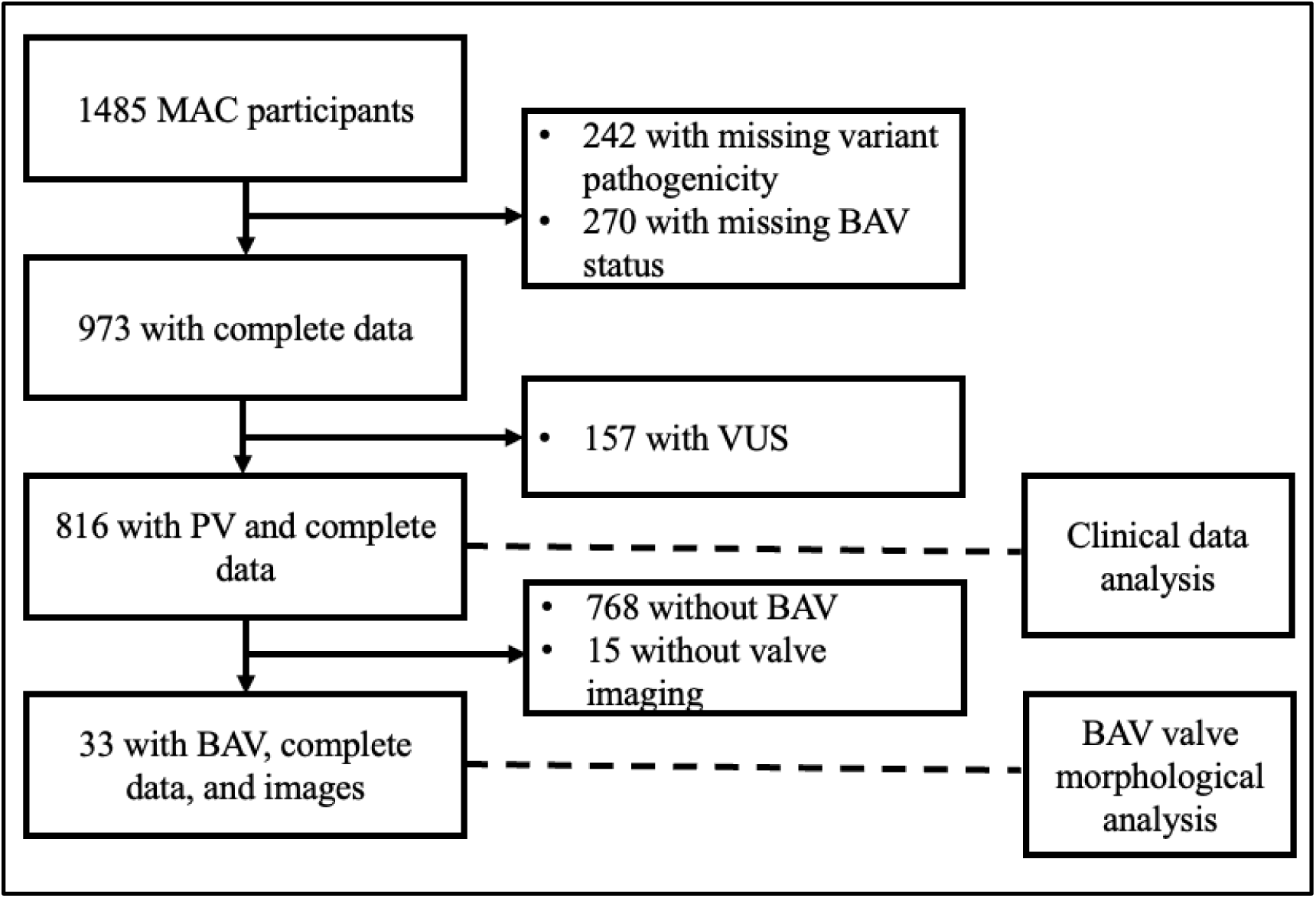
Flow chart illustrating selection process for the study. MAC: Montalcino Aortic Consortium registry; PV: pathogenic variant; BAV; bicuspid aortic valve; VUS: variant of uncertain significance.

### Clinical Features

The overall prevalence of BAV in the MAC cohort was 6% and was further increased in MAC participants with *TGFBR1* and *TGFBR2* PVs. Males with *TGFBR2* PV had the highest prevalence (12%, *P<* 0.05, Figure 2, Table S2). Notably, the difference in BAV prevalence between males and females was not statistically significant in the entire MAC cohort (29/398, 7% vs. 19/418, 5% *P=*0.10 Table 1, Figure S1) or in MAC participants with TGF-β PV.

**Figure 2.**
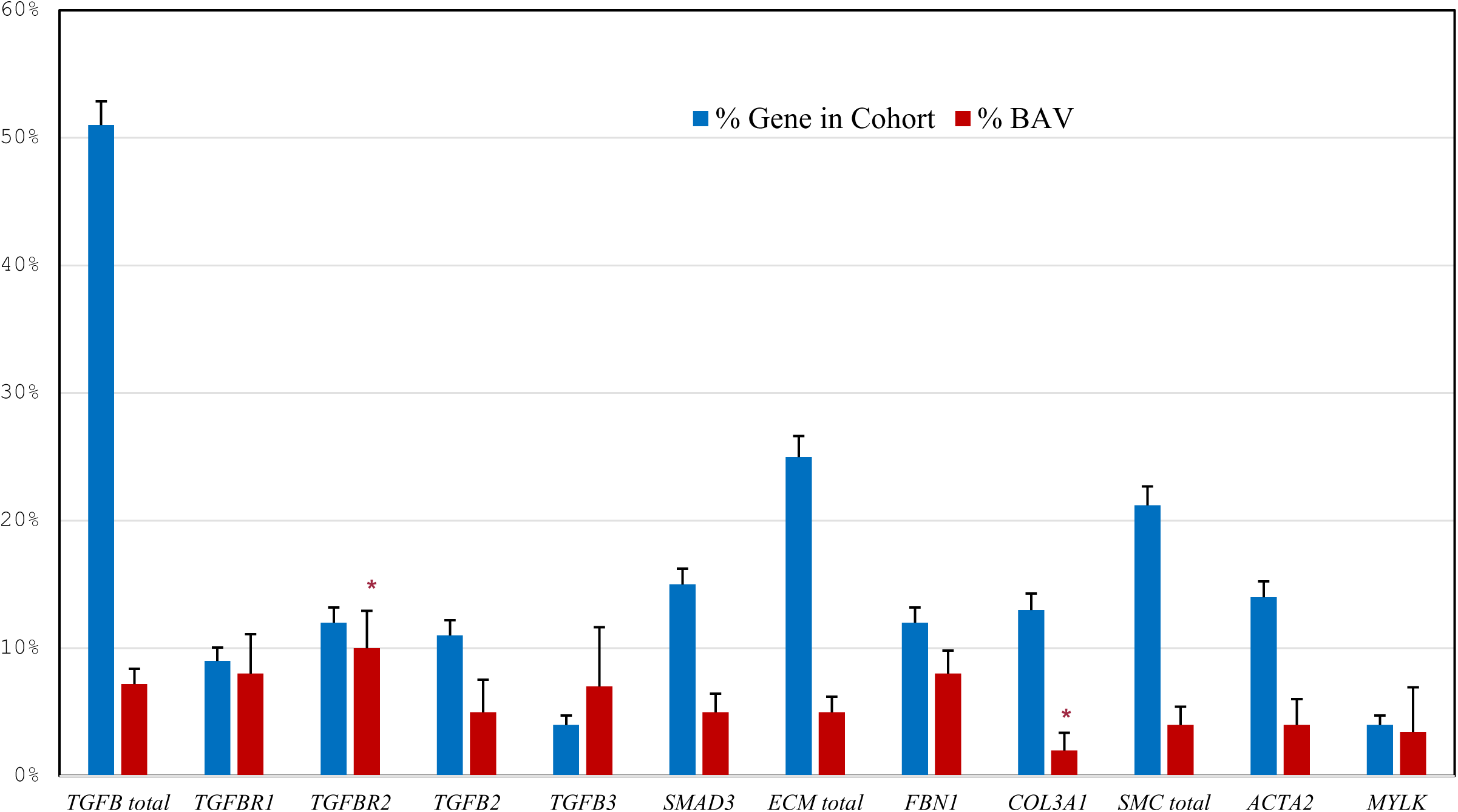
Prevalence of BAV by Gene in MAC Participants. Asterisk: p<0.05 compared to % BAV in entire cohort. n: total number of cases; %: percentage of cohort; TGF-β: Transforming growth factor-beta genes; ECM: Extracellular matrix genes; SMC: Smooth muscle cell genes; BAV: Bicuspid Aortic Valve. Genes with less than 10 individuals were excluded.

**Table 1:** Characteristics of Study Cohort.

|  | **TAV (n=768)** | **BAV (n=48)** | ***P*** |
| --- | --- | --- | --- |
| **Age at enrollment, y (IQR)** | 39 (21,52) | 29 (20,41) | <0.05 |
| **Age at first aortic event, y (IQR)** | 41 (31,51) | 25 (14,40) | <0.001 |
| **Female (%)** | 399 (52) | 19 (40) | 0.10 |
| **Weight, kg (IQR)** | 73 (59, 94) | 71 (53, 87) | 0.85 |
| **Height, cm (IQR)** | 173 (160,183) | 172 (161,180) | 0.77 |
| **TGF-β pathway variants (%)** | 387 (50) | 30 (65) | 0.14 |
| **Family history of aneurysm or dissection** | 169 (22) | 17 (35) | <0.05 |
| **Family history of PV** | 172 (22) | 3 (6) | <0.01 |
| **MVP (%)** | 66 (9) | 2 (4) | 0.42 |
| **MR (%)** | 107 (14) | 10 (21) | 0.20 |
| **Moderate or severe MR (%)** | 19 (2) | 0 (0) | 0.62 |
| **AR (%)** | 85 (11) | 18 (38) | <0.001 |
| **Moderate or severe AR (%)** | 19 (2) | 5 (11) | <0.05 |
| **AS (%)** | 8 (1) | 5 (10) | <0.001 |
| **Arrhythmia (%)** | 49 (6) | 8 (17) | <0.05 |
| **Proximal aortic repair (%)** | 227 (30) | 27 (56) | <0.001 |
| **Aortic valve surgery (%)** | 171 (22) | 25 (52) | <0.0001 |
| **Aortic dissection (%)** | 137 (18) | 6 (13) | 0.44 |
| **CHF (%)** | 15 (2) | 2 (4) | 0.26 |
| **Smoker (%)** | 134 (17%) | 6 (13%) | 0.44 |
| **Hypertension (%)** | 211 (27%) | 10 (21%) | 0.40 |
| **Pregnancy (%)** | 153 (20%) | 9 (19%) | 1.0 |

Data shown as median (Quartile 1, Quartile 3) or n (%). MVP: Any degree of mitral valve prolapse, MR: mitral regurgitation, AR: aortic regurgitation, AS: aortic stenosis, CHF: congestive heart failure, IQR: Interquartile Range; TAV: tricuspid aortic valve; PV: pathogenic variant.

The presence of any degree of AR (mild, moderate, or severe, 18/48, 38% vs. 85/768, 11%, PR 3.39 [2.24-5.14], *P<*0.001) and AS (5/48, 10% vs. 8/768, 1%, PR 10.0 [3.40, 29.41], *P<*0.001) was significantly increased in individuals with BAV. In 33 individuals with BAV who had available images, the most common valve configurations were right-left (27/33, 82%) and right-non (4/33, 12%) cusp fusion. AR was mild, moderate, or severe in 13 of BAV cases with available images (10/26 with right-left cusp fusion, Table S3), but the sample was not sufficient to distinguish associations between valve morphology and AR. Mitral valve diseases, including MR or MVP (2/48, 4% vs. 66/768, 9%, *P=*0.42), were not increased in MAC participants with BAV (Table 1). There were no reported cases of endocarditis.

Aortic events were increased in MAC participants who had HTAD PVs and also had BAV. The incidence of elective proximal aortic aneurysm repair (27/48, 56% vs. 227/768, 30%, PR 1.90 [1.45, 2.50], *P<*0.001) and aortic valve surgery (25/48, 52% vs. 171/768, 22%, PR 2.34 [1.63, 3.33], *P<*0.001) were significantly increased in the BAV group (Table 1). Symptomatic arrhythmias, including PACs, PVCs, SVTs, VT, and atrial fibrillation, were also more prevalent in participants with BAV (8/48, 17% vs. 49/768, 6%, PR 2.76 [1.24, 6.15], *P<*0.02). There were no differences in incidence of aortic dissection (6/48, 13% vs. 137/768, 18%, *P=*0.40) or CHF (2/48, 4%, vs. 15/768, 2%, *P=*0.60).

Significant aortic dilation (Z>3) at the sinuses of Valsalva (25/48, 52% vs. 128/768, 17%, PR 3.14 [2.12, 4.66], *P<*0.001) and ascending aorta (Z>3) (12/48, 25%, vs. 26/768, 3%, PR 8.19 [4.23, 15.87], *P<*0.001) was more common in MAC participants with HTAD PVs and BAV (Table 2). The pattern of aortic dilation also varied depending on BAV status. In the group with structurally normal aortic valves (TAV), isolated dilation of the sinuses (141/179, 79% vs. 16/30, 53%, PR 1.49 [1.09, 2.03], *P<*0.05) was more frequent. In the group with BAV, diffuse dilation of the sinuses and ascending aorta was more frequent (10/30, 33% vs. 20/179, 10%, PR 3.00 [1.54, 5.84], *P=*0.1). There was no significant difference in isolated ascending aortic dilation (4/30, 13% vs. 16/179, 9%, *P=*0.5, Table 2).

**Table 2:**
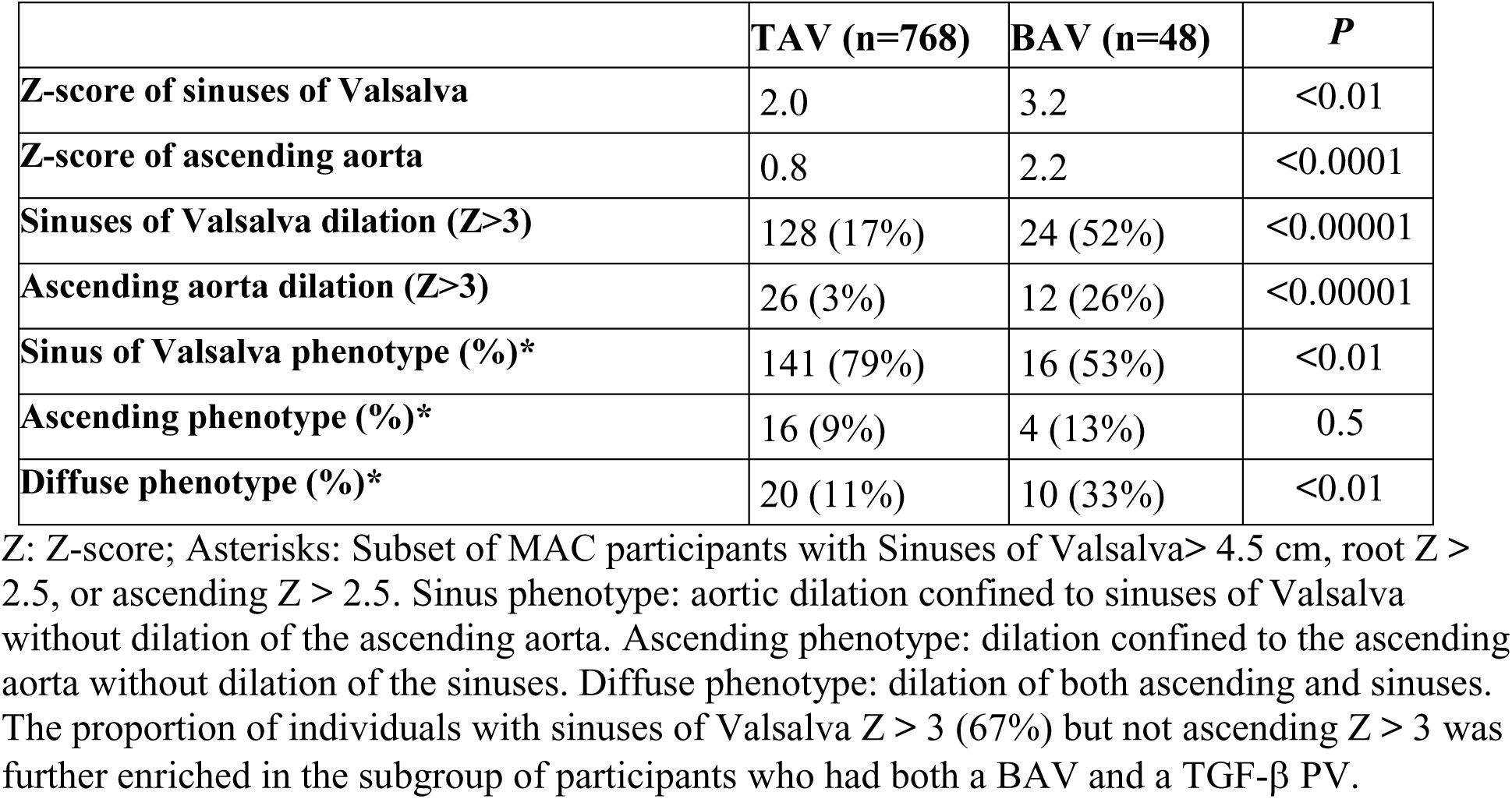
Aortic Phenotypes of Study Cohort.

Demographic comparisons showed that BAV participants were significantly younger than TAV participants (Table 1). The median age of MAC participants with BAV who experienced an aortic event was 24 years compared to 40 years for all other MAC participants. These results did not change after adjustment for baseline aortic diameter and confirmed that BAV (*P<*0.001) is an independent predictor of the composite outcome. When aortic dissection and aortic repair were considered separately, BAV remained an independent predictor of aortic repair (*P<*0.001) but not dissection. In fact, MAC participants with BAV were less likely to experience a dissection as their initial aortic event than MAC participants with TAV (137/265, 52% vs. 6/26, 23%, *P<*0.01).

### Analysis by Genotype

Half of the study cohort (417/816, 51%) had PVs in TGF-β pathway genes. The overall incidence of BAV was not significantly increased in MAC participants with TGF-β PVs compared to PVs in other genes (30/417, 7% vs. 18/399, 5%, p = 0.14 Table 3, Table S2). However, BAV was enriched in MAC participants with *TGFBR1* or *TGFBR2* PVs (16/168, 10% vs. 18/399, 5%, PR 2.11 [1.10, 4.04], *P<*0.05, Figure 2). The prevalence, types, and distribution of coding PVs in the three genes that were overrepresented in the BAV group, *TGFBR1*, *TGFBR2*, and *TGFB2*, were not significantly different in BAV and TAV cases (Figure S2). In the subgroup of MAC participants who had VUSs, the median REVEL scores of missense variants were also not significantly different between BAV and TAV cases (0.72 [0.57, 0.93] vs. 0.83 [0.57, 0.92], *P=*0.36, Figure S3).

**Table 3:** Characteristics of MAC Participants by TGF-β PV.

|  | **TGF-β PV (n=417)** | **Other PV (n=399)** | **PR (95% CI)** | ***P*** |
| --- | --- | --- | --- | --- |
| **Age** | 36 (20,52)* | 39 (22,52) | - | 0.36 |
| **BAV** | 30 (7) | 18 (5) | 1.59 (0.90-2.81) | 0.14 |
| **MVP** | 49 (12) | 19 (5) | 2.47 (1.48-4.12) | <0.001 |
| **MR** | 72 (17) | 45 (11) | 1.53 (1.08-2.16) | <0.05 |
| **Moderate or severe MR** | 8 (2) | 12 (3) | 0.64 (0.26-1.54) | 0.37 |
| **AR** | 62 (15) | 41 (10) | 1.45 (1.00-2.08) | 0.06 |
| **Moderate or severe AR** | 9 (2) | 10 (3) | 0.86 (0.35-2.10) | 0.82 |
| **AS** | 8 (2) | 5 (1) | 1.53 (0.51-4.64) | 0.58 |
| **Arrhythmia** | 34 (8) | 23 (6) | 1.41 (0.85-2.36) | 0.22 |
| **Proximal aortic repair** | 140 (34) | 114 (29) | 1.18 (0.96-1.44) | 0.13 |
| **Aortic valve surgery** | 127 (31) | 69 (17) | 1.76 (1.36-2.28) | <0.0001 |
| **Aortic dissection** | 55 (13) | 88 (22) | 0.60 (0.44-0.81) | <0.001 |
| **CHF** | 11 (3) | 6 (2) | 1.75 (0.65-4.70) | 0.33 |

Asterisk: Median (Quartile 1, Quartile 3). BAV: Bicuspid aortic valve. MVP: Any degree of mitral valve prolapse, MR: mitral regurgitation, AR: aortic regurgitation, AS: aortic stenosis, CHF: congestive heart failure, TGF-β : transforming growth factor-beta, PR: prevalence ratio, CI: Confidence Interval.

MR (72/417, 17% vs. 45/399, 11%, PR 2.47 [1.48, 4.12], *P<*0.001) and MVP (49/417, 12% vs. 19/399, 5%, PR 2.47 [1.48, 4.12], *P<*0.001) were enriched in MAC participants with TGF-β PVs compared to other PVs (Table 3). AR (62/417, 15% vs. 41/399, 10%, *P=*0.06, Figure S4) and AS (8/417, 2% vs. 5/399, 1%, *P=*0.58) were not significantly increased in the TGF-β group.

Elective aortic valve surgeries were increased in MAC participants with TGF-β PVs compared to other PVs (110/387, 28% vs. 69/399, 17%, PR 1.64 [1.26-2.15], *P<*0.0001) and were further enriched in participants with BAV and TGF-β PVs (17/30, 57%, vs. 110/387, 28%, PR 1.99 [1.40-2.83], *P<*0.0001), who were also more likely to undergo elective aortic repair (18/30, 60% vs. 122/387, 32%, PR 1.90 [1.37-2.64], *P<*0.001, Figure 3, Figure S7). Arrhythmias (6/30, 20% vs. 28/387, 7%, PR 2.76 [1.24, 6.15] *P<*0.05) and AR (13/30, 43% vs. 49/387, 13%, PR 3.42 [2.11, 5.56], *P<*0.001) were significantly enriched in this subgroup, but CHF (1/30, 3% vs. 10/387, 3%) and aortic dissection (2/30, 7%, vs. 53/387, 14%, *P=*0.40) were rare and were not increased.

**Figure 3:**
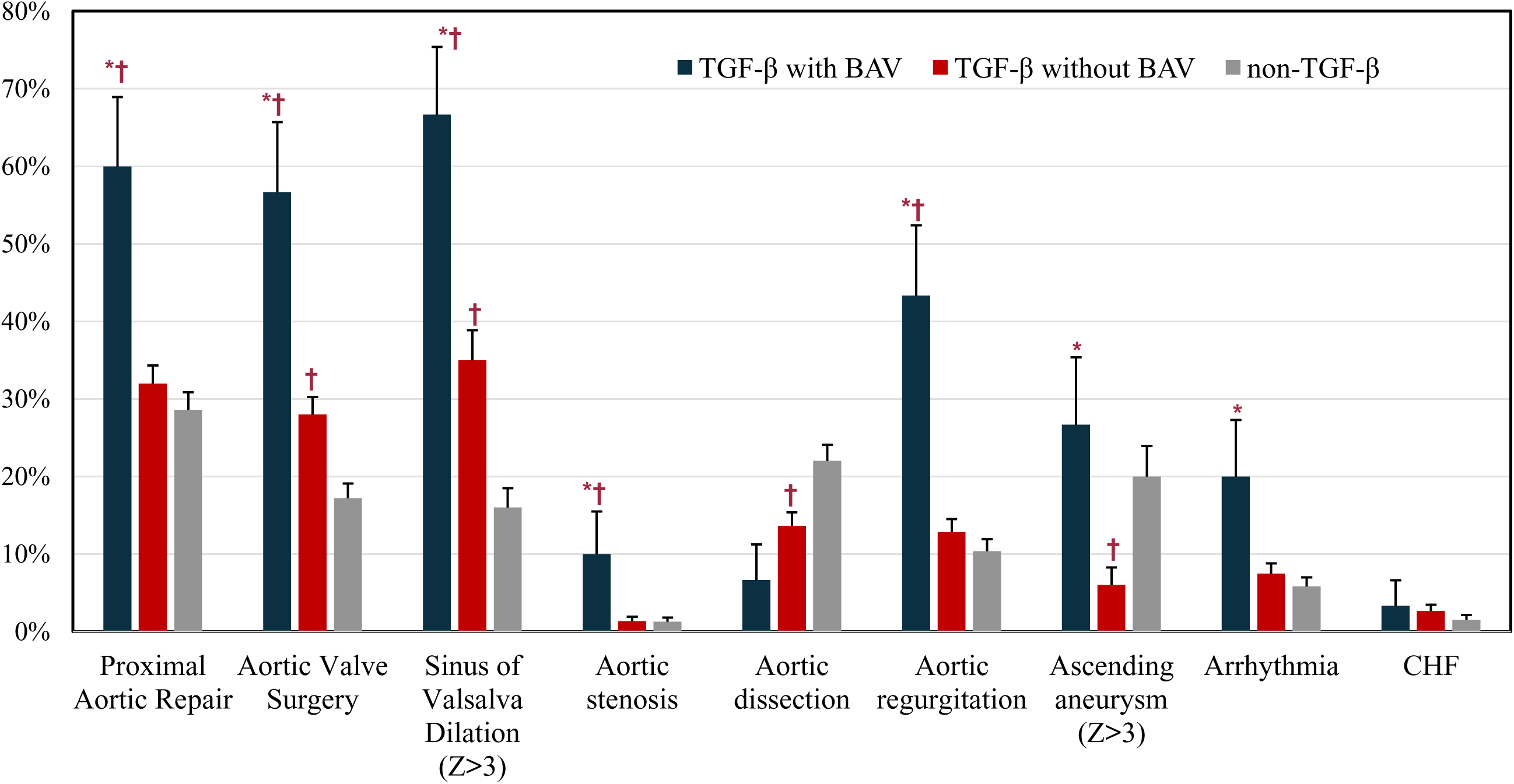
Aortic Outcomes by TGF-β PV and BAV. Asterisk: p<0.05 compared to ‘TGF-β without BAV’ group. Dagger: p<0.05 compared to ‘non-TGF-β’ group. Aortic endpoints were enriched in the TGF-β group and further enriched in MAC participants who had TGF-β PV and BAV.

Significant aortic dilation at the sinuses of Valsalva (Z>3) was increased in participants with TGF-β PVs (91/258, 35% vs. 42/258, 16%, PR 2.17 [1.57-2.99], *P<*0.0001) and was further increased if they also had a BAV (20/29, 69% vs. 91/258, 35%, PR 1.96 [1.40-2.63], *P<*0.0001) (Figure 3). At the time of elective aortic operations, the median sinus and ascending Z-scores were not statistically different for the BAV and TAV groups (Figure S5). Individuals with BAV and TGF-β PVs were also more likely to be diagnosed with an ascending aortic aneurysm (8/28, 29% vs. 9/156, 5%, PR 4.95 [2.09, 11.74], *P<*0.01). The age when the initial aortic event occurred (38.89 ± 15.89 years vs. 39.42 ± 16.05 years, t(286) = 0.28, *P=*0.8) was not different for individuals with TGF-β PVs compared to other MAC participants. Initial aortic events occurred much earlier in participants who had both TGF-β PVs and BAV compared to participants who had TGF-β PVs but did not have BAV (25.06 ± 18.6 years vs. 40.6 ± 14.7 years, t(142) = 3.86, *P<*0.001) and was comparable to participants with *FBN1* PVs (25.06 ± 18.62 years vs. 23.61 ± 14.64 years, t(37) = 5.33, *P*=0.79).

The overall prevalence of BAV was significantly higher in MAC participants with VUSs than in MAC participants with PVs (16/157, 10%, vs. 48/816, 6%, PR 1.73 [1.01-2.97], *P<*0.05) (Figure S6). No single gene was significantly enriched in the VUS subgroup (Figure S7). When individuals with VUSs were included in the analysis, aortic outcomes did not change, with more frequent proximal aortic repair (36/64, 56% vs. 268/907, 30%, PR 1.90 [1.50-2.42], *P<*0.001), aortic valve surgery (33/64, 52% vs. 198/907, 22%, PR 2.36 [1.81-3.09], *P<*0.0001), and AR (22/64, 34% vs. 110/907, 12%, PR 2.83 [1.94-4.15], *P<*0.001) in the BAV group but no increase in aortic dissection (9/64, 14% vs. 167/907, 18%, PR 0.76 [0.41-1.42], *P=*0.41]).

## Discussion

In an analysis of MAC registry participants, we found that BAV is enriched up to ten times its population prevalence, is highest in cases with *TGFBR1* and *TGFBR2* PVs, and is associated with accelerated aortic disease, leading to earlier and more frequent elective proximal thoracic aortic repair. The lifetime risk of a thoracic aortic aneurysm is as high as 70% in BAV natural history studies [5,6]. Altered hemodynamics due to BAV can produce asymmetric shear stress in the ascending aorta driving the progression of ascending aortic dilation and may help explain why BAV is predominantly associated with ascending aortic aneurysms [26]. In contrast, Marfan syndrome (due to *FBN1* PVs) and other heritable aortopathies predominantly affect the sinuses of Valsalva with secondary dilation of the proximal ascending aorta as the disease progresses. Notably, the risk for aortic dissection in isolated BAV cohorts is substantially lower than that observed in cases with HTAD, even after accounting for aortic size [6,27]. How BAV may interact with an HTAD PV to modify the trajectory of aortic enlargement or change dissection risk is a critical unresolved determinant of prognosis.

We confirmed these trends in MAC by showing that participants with BAV developed more extensive dilation of the ascending aorta and that BAV was an independent predictor of elective proximal thoracic aortic repair. Individuals with BAV experienced their initial aortic event at significantly younger ages (24 vs. 40 years) than other MAC participants, primarily due to elective repair of rapidly enlarging aortic aneurysms. Correspondingly, moderate or severe AR due to progressive ascending dilation with effacement of the sinotubular junction was significantly more frequent in MAC participants with BAV and was the predominant driver of early presentations due to valve disease. MAC participants with BAV reached aortic diameter thresholds for preventative surgery earlier but did not experience increased rates of aortic dissection, although the median aortic diameters of the BAV and TAV groups were equivalent at the time of elective repair (Figure S8). Our data suggest that BAV does not interact with HTAD PVs to increase dissection risk but may be associated with a protective effect against dissection that will require confirmation in larger cohorts with longer follow-up. Notably, a similar effect of BAV on increased dilation and reduced dissection rates was observed in an *FBN1* cohort with Marfan syndrome [28].

These findings have implications for the role of genetic testing in selected individuals with BAV, especially those who present due to early onset valve or aortic complications and have a family history of thoracic aortic disease or syndromic features suggestive of LDS. A small proportion of young individuals with these same characteristics was independently identified in adult BAV cohorts [29]. While aortic dissection rates associated with BAV are generally low, this ‘complex valvulo-aortopathy’ BAV phenotype with large thoracic aortic aneurysms or severe aortic valve disease before age 30, syndromic features, or complex congenital heart disease was found to have a higher adjusted mortality rate [3]. The MAC BAV cohort exhibits complex aortopathy features and includes cases in which BAV drove the initial presentation and diagnosis of some individuals with HTAD PVs. Genetic data can change the clinical management of these cases because there are lower gene-based thresholds for elective repair of aneurysms if an HTAD PV is identified [18]. Earlier recognition of individuals with BAV resulting in more frequent preventative aortic repair may also decrease dissection rates [30]. These findings underscore the importance of prompt diagnosis and individualized management of BAV disease associated with HTAD PVs.

The prevalence of BAV was highest in MAC participants with PVs in TGF-β genes, especially *TGFBR1* and *TGFBR2*, with a similar relative enrichment as in previous observations [31]. TGF-β genes are required for fetal left ventricular outflow tract development and valve morphogenesis and altered TGF-β signaling during embryogenesis can cause BAV [32,33]. In the MAC cohort, the highest diagnostic rates of TAA leading to elective aortic repair and valvular heart disease occurred in MAC participants with both a TGF-β PV and a BAV. These findings illustrate how the combination of an HTAD PV disrupting aortic function and altered hemodynamics due to BAV may accelerate the presentation of aortic disease.

There were noteworthy patterns in the gene and sex-based analysis of BAV. *SMAD3* PVs are associated with prominent mitral valve pathologies, including mitral valve prolapse and mitral annular disjunction [34]. In this study, we observed that BAV was not enriched in MAC participants with *SMAD3* PVs, in contrast to other TGF-β genes that were associated with higher BAV rates but relatively fewer mitral phenotypes. Valve phenotypes along with other factors such as the pattern of branch vessel involvement may be useful to distinguish LDS subtypes and are reflective of distinct roles for the different HTAD TGF-β genes during heart development [35]. There were no significant correlations between BAV and the type or location of variants in *TGFBR1*, *TGFBR2*, or *TGFB2*, which had the most BAV-associated PVs. The anatomic configurations of the valve cusps were similar to the distribution of BAV variants in the general population (Table S3). In an analysis of recurrent PVs in HTAD TGF-β genes, the penetrance of BAV was low (7-50%, Table S4).

In the GenTAC cohort, our early onset BAV with complications cohort, and the Olmstead County BAV cohort, the male to female ratio ranges between 2 and 3, i.e., 70-78% male [8,36,37]. In MAC, the male to female ratio of BAV participants who had an HTAD VUS was also 2-3 but was attenuated in participants with an HTAD PV (1.6) and did not vary according to the mutated gene. Overrepresentation of women in the MAC BAV cohort may be explained by study design and recruitment differences. It is also possible that the presence of a BAV may increase the clinical recognition of individuals with HTAD PVs due to exacerbation of valve or aortic disease. Our data fit a model in which disease penetrance and severity are determined by interactions between multiple genetic variants and modifiable risk factors such as BAV, smoking, pregnancy, and hypertension, which can accelerate the onset and increase the severity of HTAD. Predictive models for HTAD should include BAV along with other clinical and genetic data for risk stratification and clinical decision making about the intensity of clinical surveillance or the timing of elective interventions.

Limitations of this study relate to the fact that MAC study sites are tertiary referral centers that recruit participants with relatively severe HTAD phenotypes. Therefore, individuals with more prominent aortic disease may be overrepresented and this could inflate our effect estimates. This study was relatively underpowered to detect genotype-phenotype correlations due to small samples and incomplete data. Future studies that include a more representative distribution of HTAD genotypes with more complete follow-up are needed to translate these observations into clinical recommendations for management of BAV and HTAD.

Taken together, these data show that BAV is enriched in patients with HTAD, especially those with TGF-β gene PVs, and can accelerate the development of clinically significant aortic disease without clearly increasing dissection risk. These findings support targeted genetic testing of patients with BAV who present with syndromic features or a family history of thoracic aortic disease. When an HTAD PV is identified in a patient with BAV, there are immediate implications for clinical management, including lower thresholds for elective repair, closer surveillance intervals, and more intensive risk factor management. We found no evidence that elective surgical repair thresholds should be lowered for HTAD patients with BAV.

## Data Availability

The data that support the findings of this study are available from the corresponding author on reasonable request. Individual-level data from the Montalcino Aortic Consortium (MAC) registry cannot be made publicly available due to participant privacy and institutional data-sharing agreements, but de-identified datasets underlying the analyses may be provided to qualified investigators upon request and pending appropriate approvals.

https://montalcinoaorticconsortium.org

## Acknowledgements

We thank the study participants and referring clinicians without whom this study would not have been possible. Special thanks to Niveya James for assistance with MAC registry data.

## Appendix

The Montalcino Aortic Consortium investigators are: Dianna M. Milewicz, MD, PhD; Reed Pyeritz, MD, PhD; Julie De Backer, MD, PhD; Catherine Boileau, PhD; Guillaume Jondeau, MD, PhD; Alan Braverman, MD; Shaine A. Morris, MD; Arturo Evangelista, MD; Maral Ouzounian, MD, PhD; Anji Yetman, MD; Richmond, Jeremy, MB, BS, PhD; Sherene Shalhub, MD, MPH; Eloisa Arbustini, MD; Lesley Ades, MD; Nanette Alvarez, MD; Anne Child MD; Bo Carlberg, MD, PhD; Ismail El-Hamamsy, MD, PhD; David Chitayat, MD; Anne De Paepe, MD, PhD; Richard B. Devereaux, MD; Bridgete Fernandez, MD; Josephine Grima, PhD; Maarten Groenink, MD, PhD; Gabrielle Horne, MD, PhD; Yskert von Kodolitsch, MD; Ronald Lacro, MD; Scott A. LeMaire, MD; Alex V. Levin, MD; David Liang, MD, PxhD; Irene Maumenee, MD; Vivienne McConnell; Rocio Moran, MD; Hiroko Morisaki, MD, PhD; Takayuki Morisaki, MD, PhD; Alex Pitcher, PhD; Nancy Poirier, MD; Francesco Ramirez, PhD; Peter Robinson, MD; Dieter Reinhardt, PhD; Meike Rybczynski, MD; George Sandor, MD; Lynn Sakai, PhD; Denver Sallee, MD; Michael N. Singh, MD; Bert Callewaert, MD, PhD; Ellen Regalado, CGC; Marion A. Hofmann-Bowman, MD, PhD; Fabien Labombarda, MD; Laurence Faivre, MD, PhD; Claire Bouleti, MD, PhD; Marjolijn Renard, PhD; Derek Human, BA, BM. BCh; Zoltan Szabolcs, MD, PhD; Andrew Michael Crean, MD; Joseph Bavaria, MD; Apostolos Psychogios, MD; Vidyasagar Kalahasti, MD; Philip Giampietro, MD, PhD; Laura Muiño-Mosquera, MD; Gisela Teixido-Tura, MD, PhD; Olivier Milleron, MD; Mark Evan Lindsay, MD, PhD; Juan Manuel Bowen, MD; Shuping Ge, MD; Anthony Caffarelli, MD; Jolien Roos- Hesselink, MD; Mary B. Sheppard, MD; Andrew Choong, MBBS; Enid Neptune, MD; Zhou Zhou, MD, PhD; Alessandro Pini, MD; Germano Melissano, MD; Elisabetta M. Mariucci, MD; Elena Cervi, MD, PhD; Melissa L. Russo, MD; Pascal-Nicoloas Bernatchez, PhD; Mitra Esfandiarei, PhD; Julien Marcadier, MD; Seda Tierney, MD; Jessica Wang, MD, PhD; Kevin Michael Harris, MD; Sara B. MacKay, CCGC; Andrea L. Rideout, CCGC; Sandhya Prakash, MD; Anthony Martin Vandersteen, PhD; Nadine Hanna, PharmD, PhD; Carine Le Goff, PhD; Quentin Pellenc, MD; Patrick Sips, PhD; Scott Michael Damrauer, MD; Lois Starr, MD, PhD; Bo Yang, MD, PhD; Jay B. Shah, MD; Justin T Tretter, MD; Irman Forghani, MD; Kelly Cox, MD; Siddharth K. Prakash, MD, PhD; Sachiko Kanki, MD, PhD; Justin A Weigand, MD; Taylor A. Beecroft, CGC; Alan Frederick Rope, MD; Yaso Emmanuel, MBChB, DPhil.; David Dichek, MD; Michelle Keir, MD; Klaus Kallenbach, MD; Talha Niaz, MBBS; Mirela Tuzovic, MD; Anthony L. Estrera, MD; Timothy Carter, MD; Martin Czerny, MD; Anna Sabaté Rotés, MD, PhD; Brett Carroll, MD; William T. Brinkman, MD; David R. Murdock, MD; Christopher Phillips Jordan, MD.

## Nonstandard abbreviations and Acronyms

HTAD: Heritable Thoracic Aortic Disease
BAV: Bicuspid Aortic Valve
PV: Pathogenic Variant
TGF-β: Transforming Growth Factor-Beta
SMC: Smooth Muscle Cell
ECM: Extracellular Matrix
VUS: Variant of Uncertain Significance

## Supplemental Material

**Table S1.**
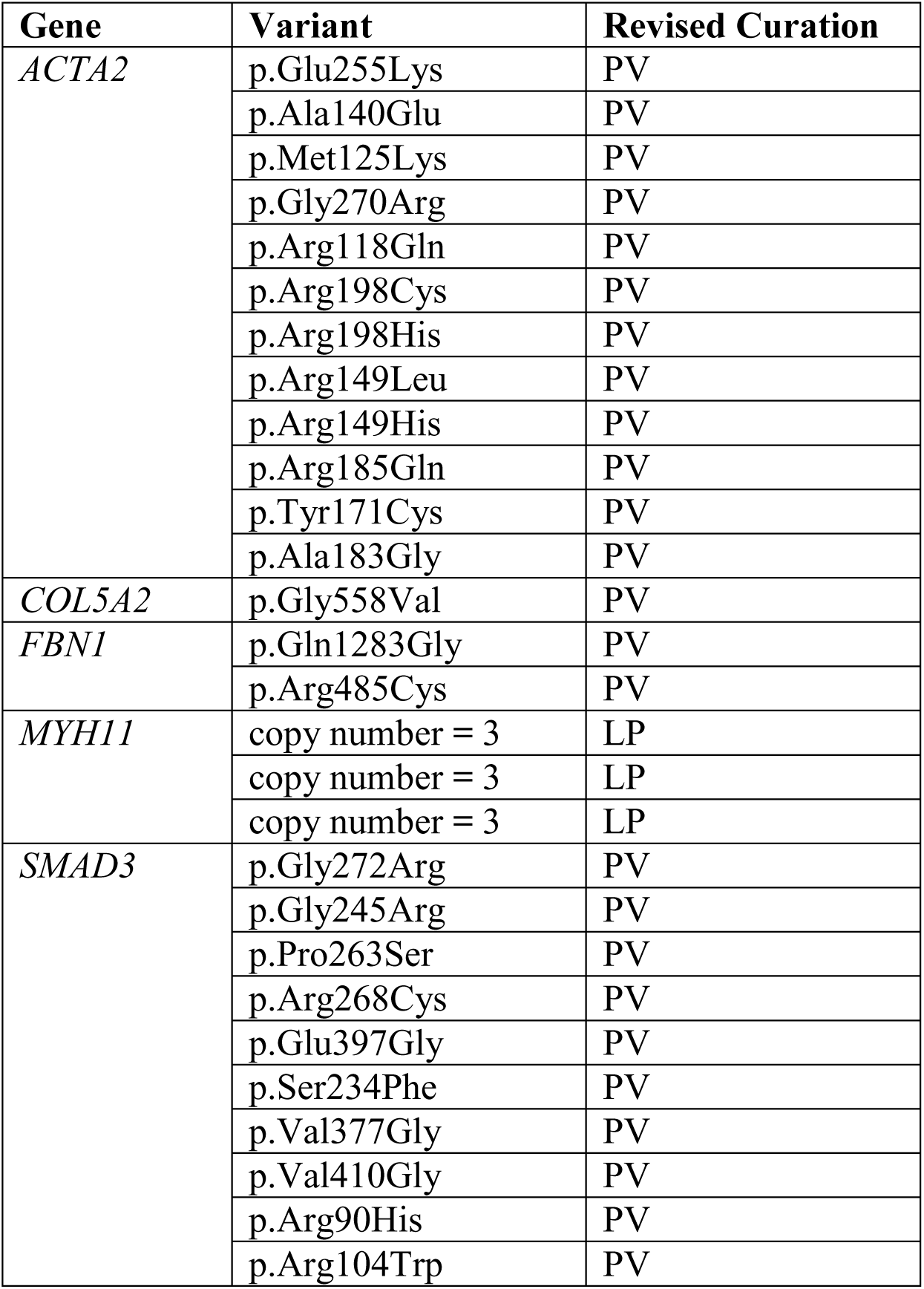
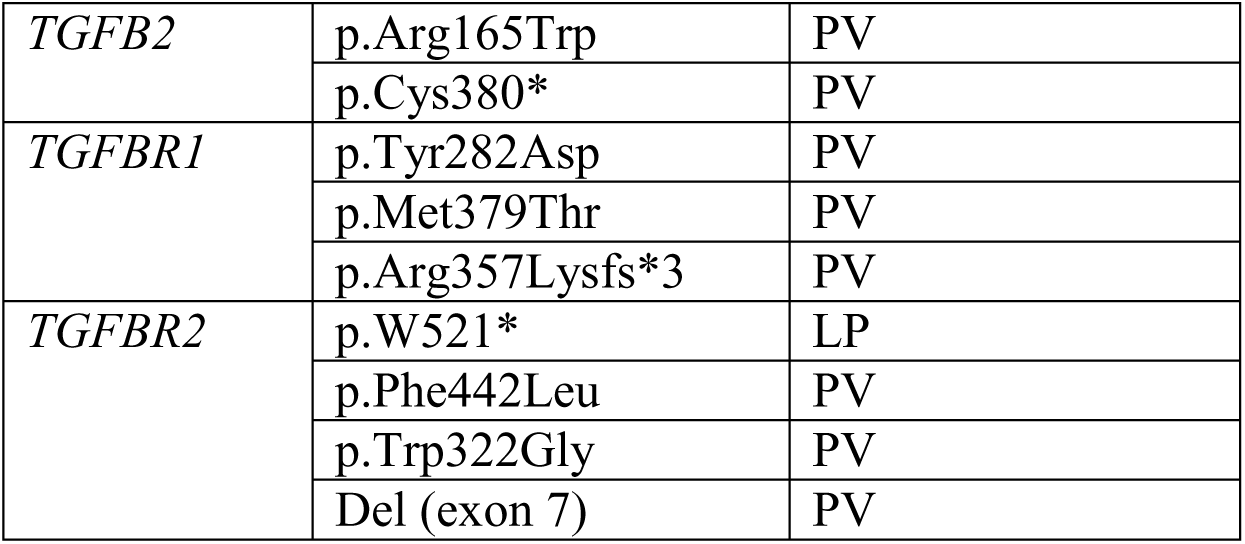
Variants of Uncertain Significance (VUSs) that were reclassified. Duplicates not shown.

**Table S2.**
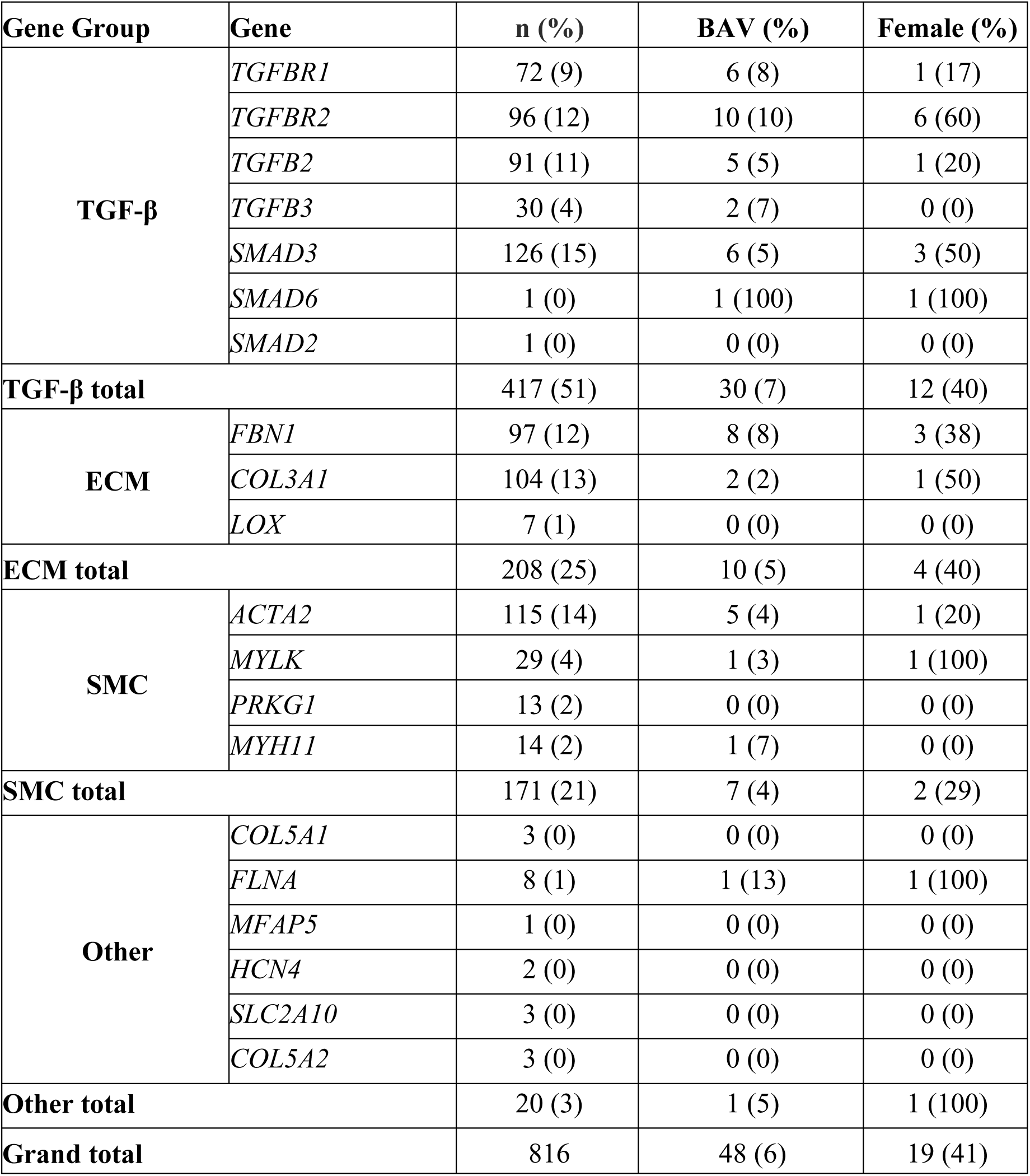
Prevalence of BAV by Gene and Sex in MAC Participants. n: total number of cases; %: percentage of cohort; TGF-β: Transforming growth factor-beta; ECM: Extracellular matrix; SMC: Smooth muscle cell; BAV: Bicuspid aortic valve.

**Table S3.**
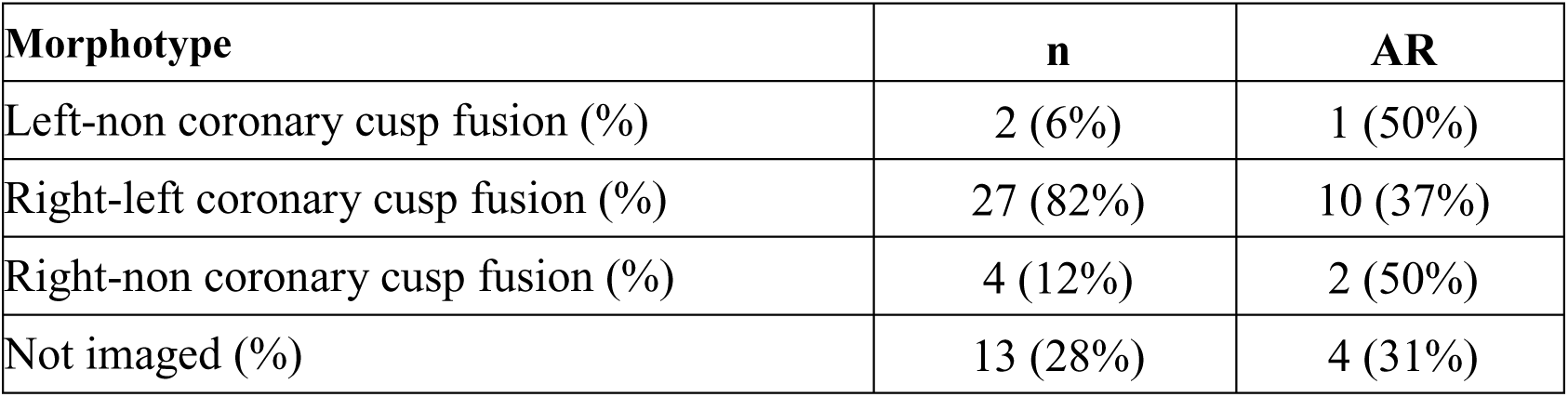
Aortic Regurgitation by BAV Morphology (n=33). AR: mild, moderate, or severe aortic regurgitation.

**Table S4.**
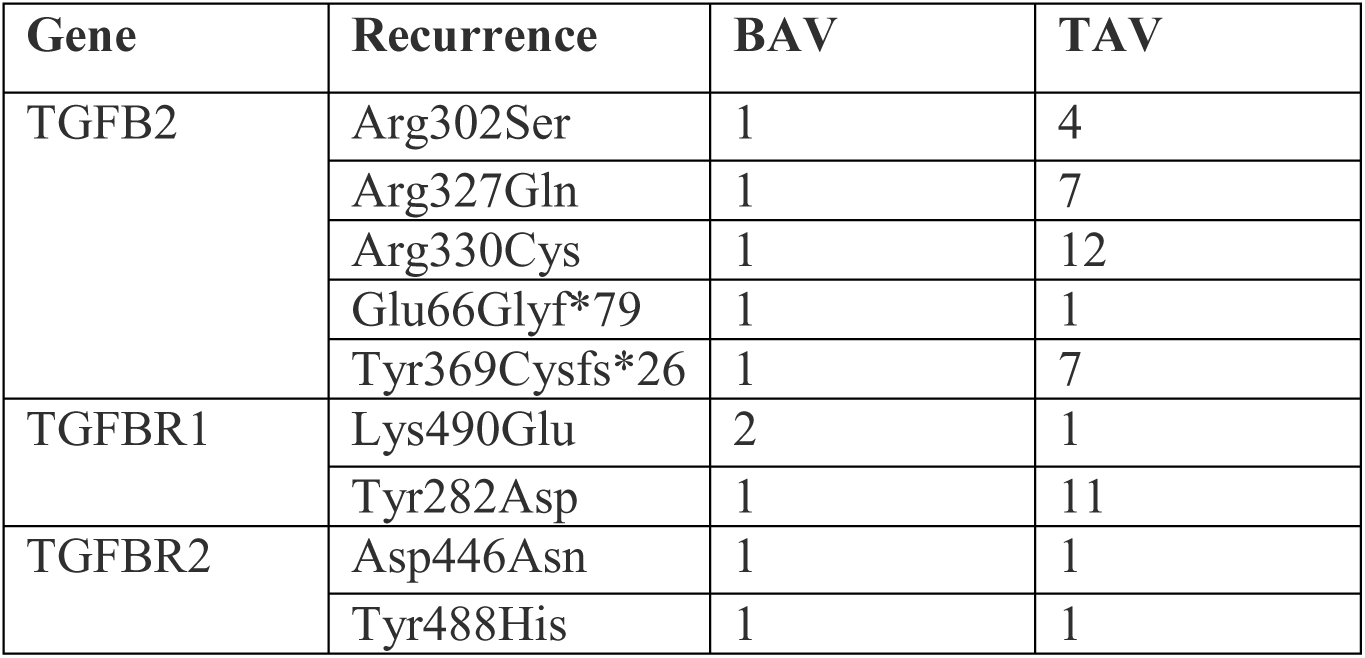
Prevalence and penetrance of BAV in MAC participants with recurrent variants in *TGFB2*, *TGFBR1*, and *TGFBR2*. Only variants with at least one BAV case are shown.

**Figure S1.**
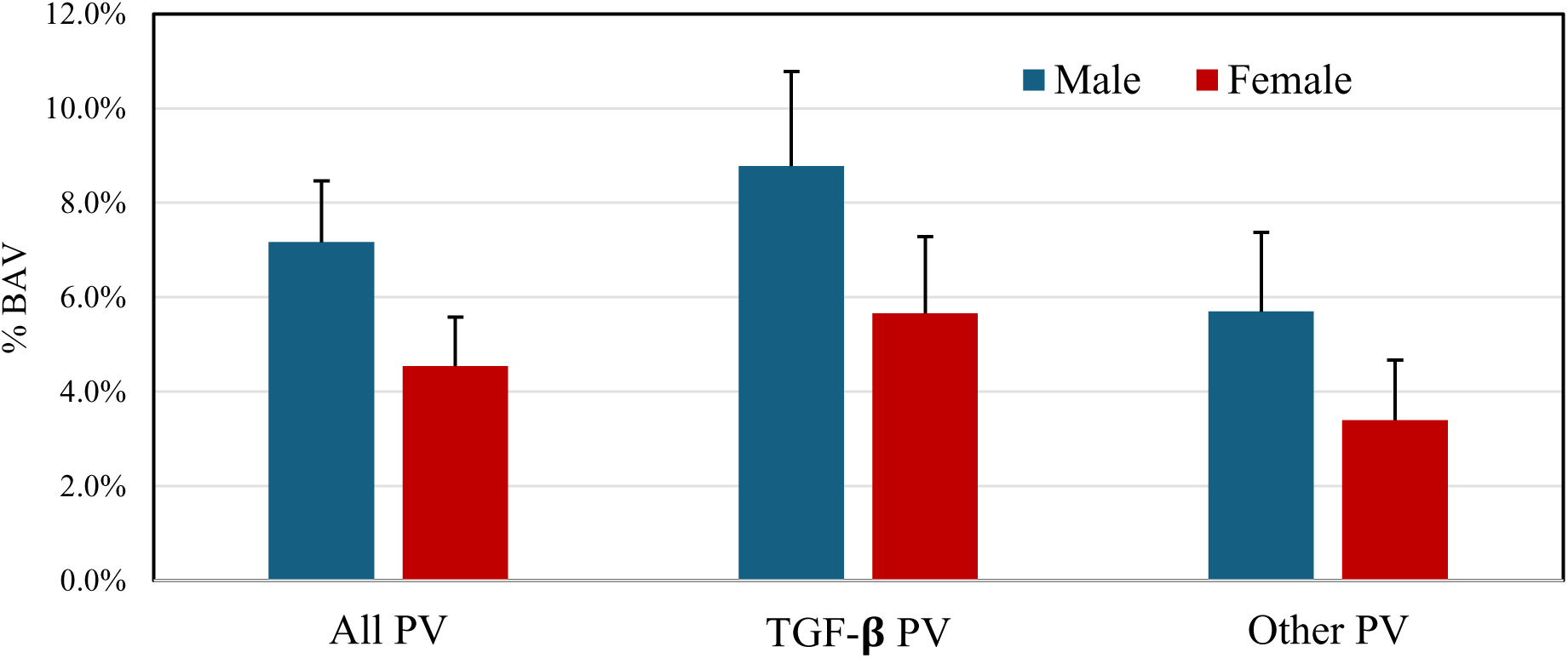
Prevalence of BAV by Sex in MAC Participants. BAV: bicuspid aortic valve. There was no difference in BAV prevalence in males compared to females in the entire cohort (p=0.13), in the group with TGF-𝛃 PVs (p=0.18), and in the group with all other PVs (p=0.46).

**Figure S2.**
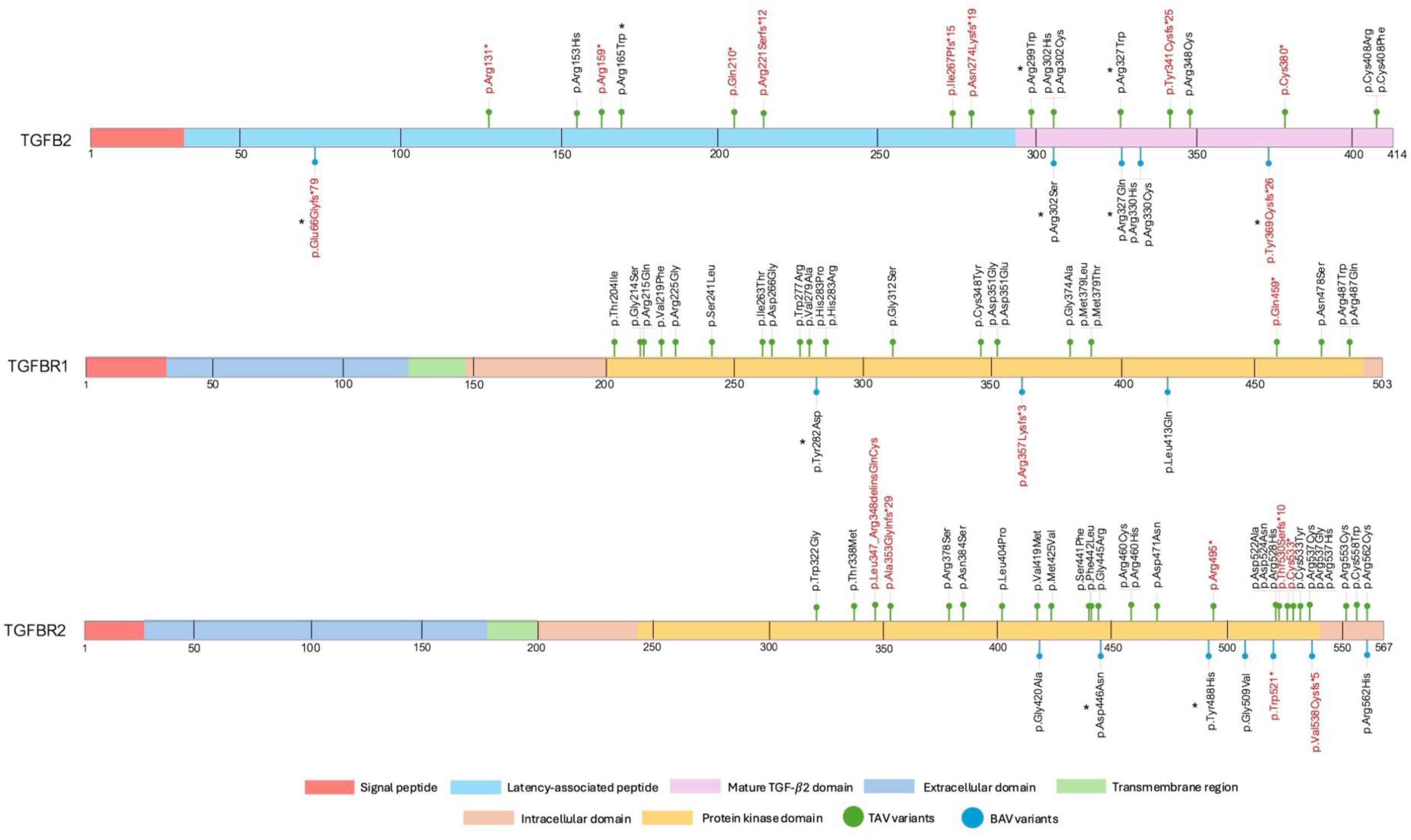
Protein maps of Tgfbr1, Tgfbr2, and Tgfb2 illustrate the location and types of coding PV in the MAC cohort by BAV status. Asterisks: recurrent variants; red text: predicted loss of function variants.

**Figure S3.**
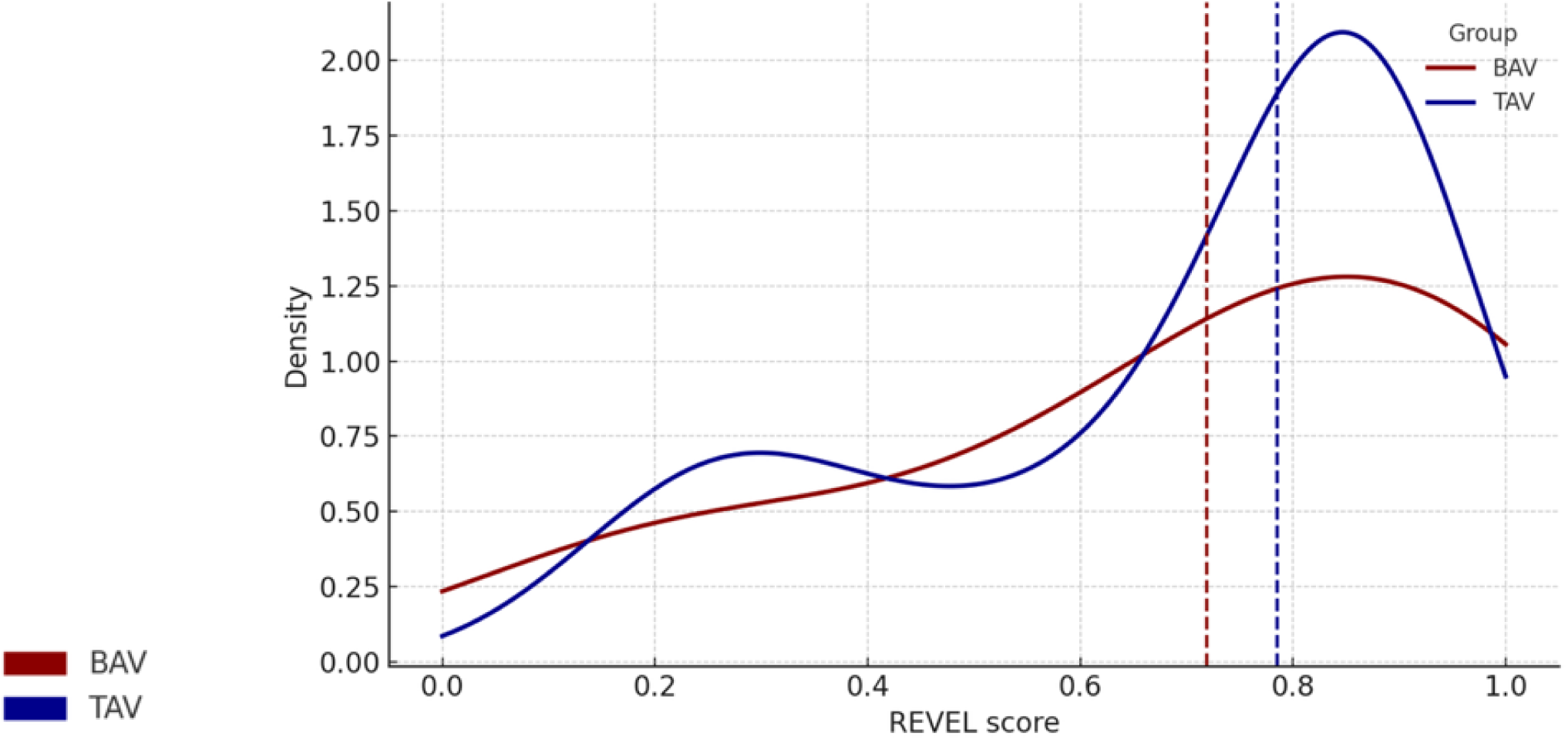
Distribution of VUS REVEL Scores For BAV and TAV MAC participants. Median REVEL scores were not significantly different.

**Figure S4.**
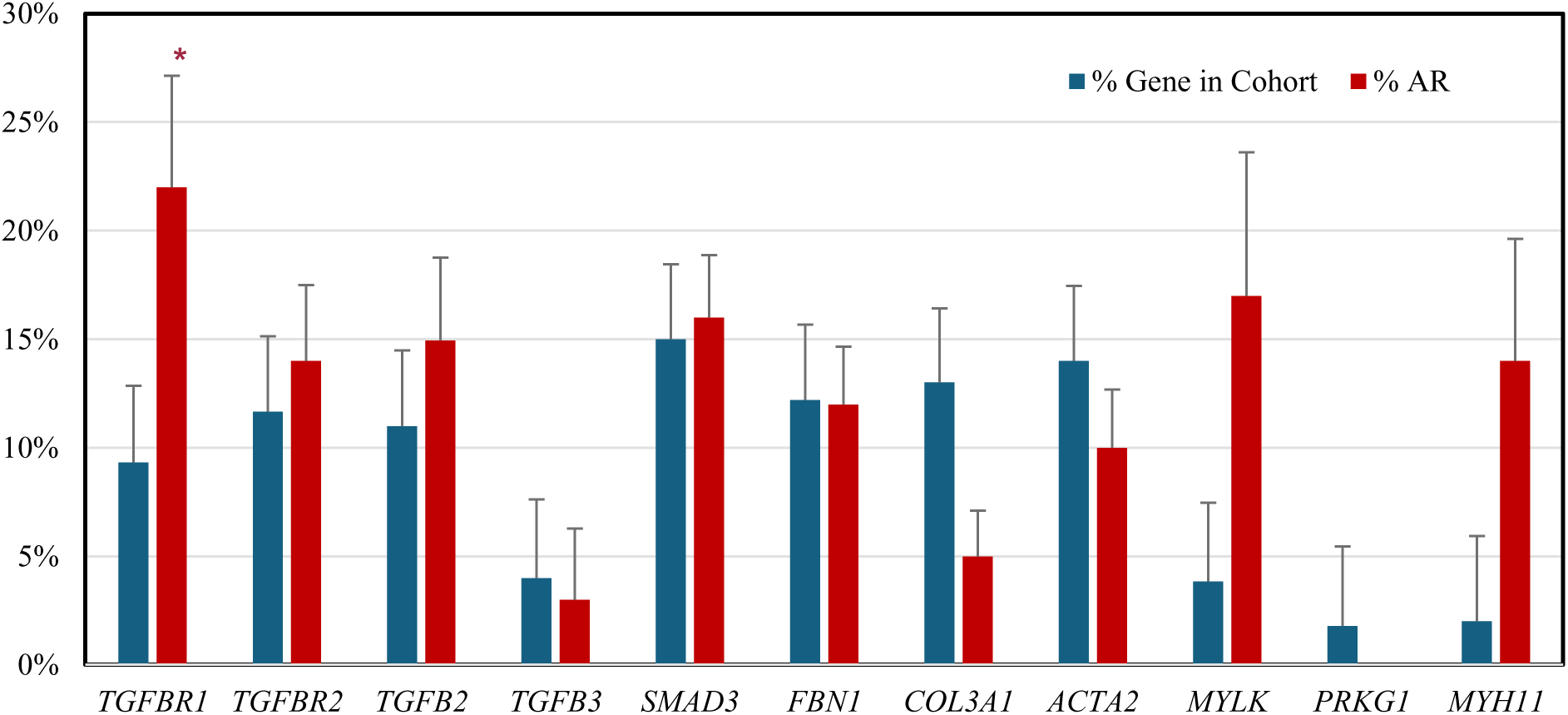
Prevalence of AR by HTAD PV. Asterisk: p<0.05 compared to % AR in other HTAD genes. AR: mild, moderate, or severe aortic regurgitation. The prevalence of AR was enriched in the group of MAC participants with TGF-β PVs but was only significant for one individual gene, *TGFBR1*. *SMAD6, SMAD2, COL5A1, MFAP5, HCN4, SLC2A10, COL5A2, LOX, FLNA* PVs were excluded due to having fewer than 10 cases.

**Figure S5.**
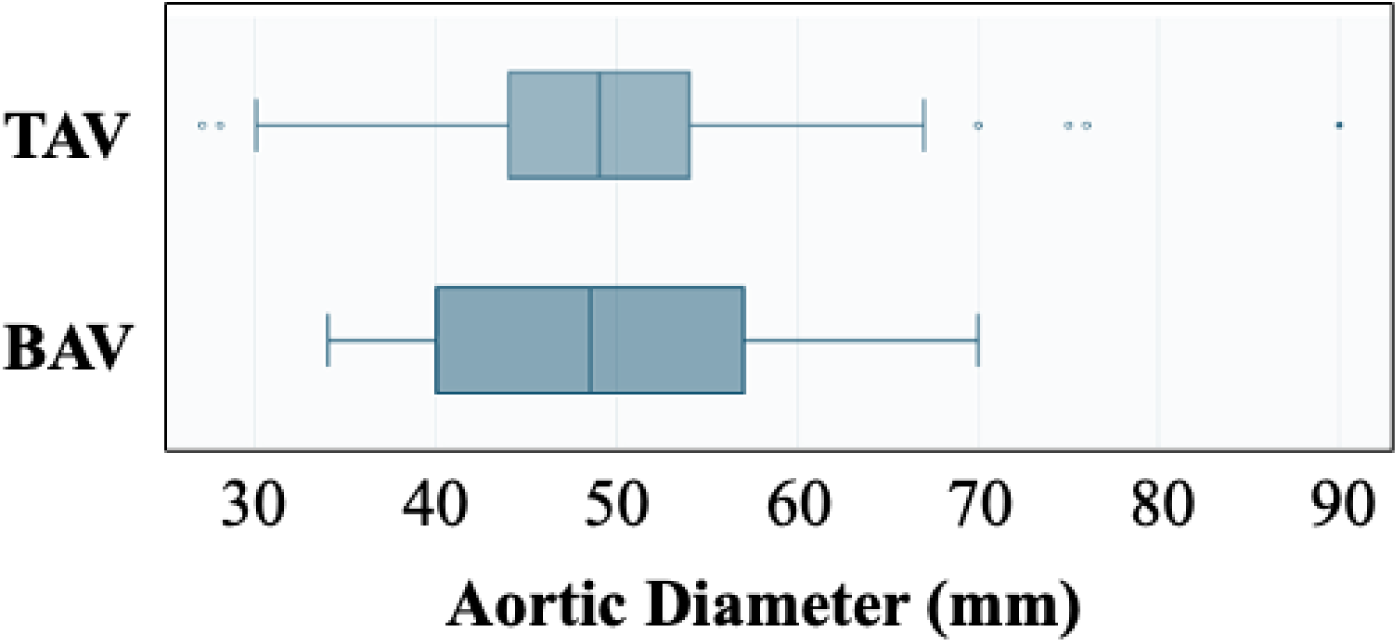
Box and whisker comparison plots of the maximum aortic diameters at the time of elective aortic repair operations for MAC participants with bicuspid (BAV) or tricuspid (TAV) aortic valves. The median diameters (vertical lines) were not significantly different (*P=*0.98).

**Figure S6.**
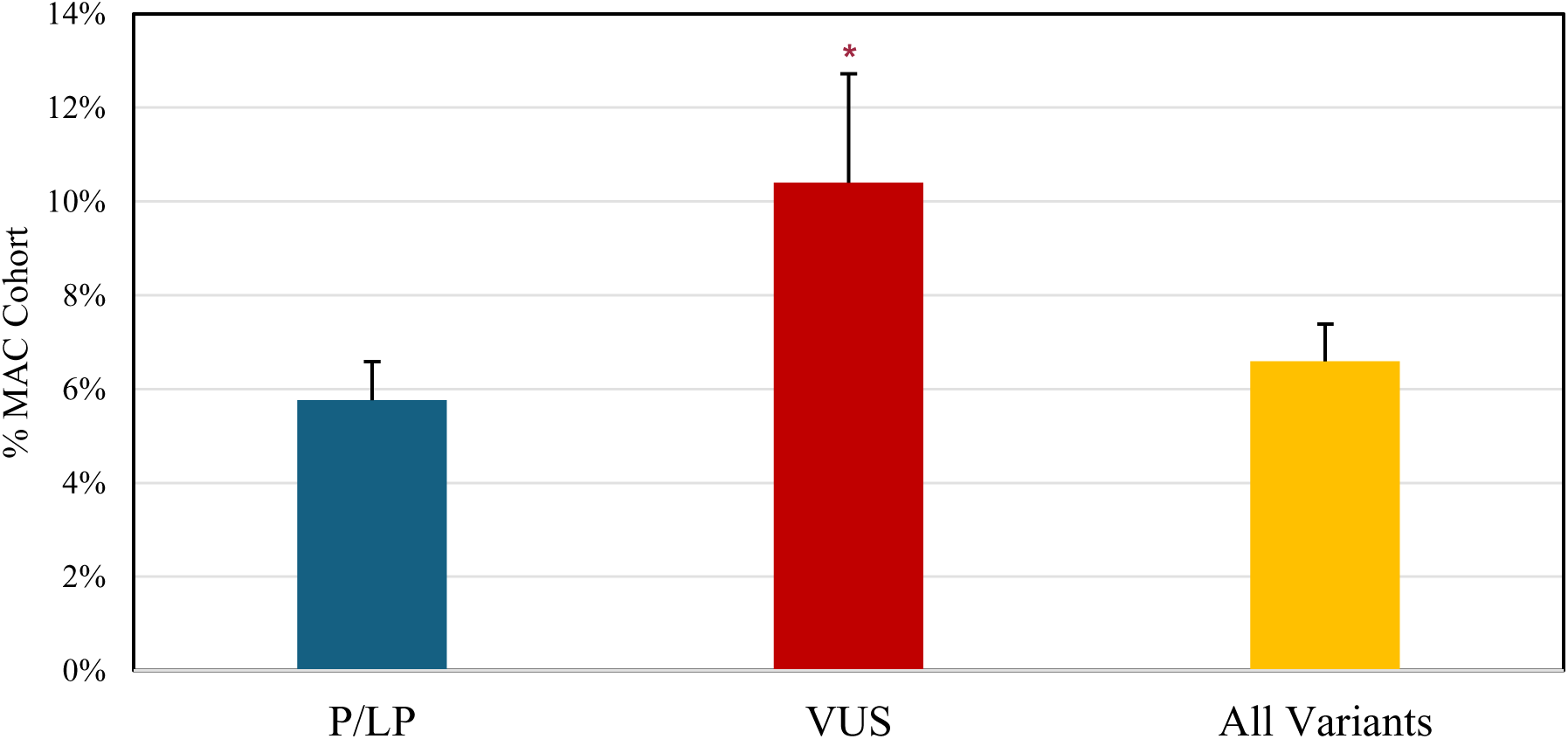
Prevalence of BAV in MAC by variant classification. All: all variants, P/LP: pathogenic and likely pathogenic variants, VUS: variants of uncertain significance, BAV: Bicuspid aortic valve. Asterisk: <0.05 compared to P/LP group.

**Figure S7.**
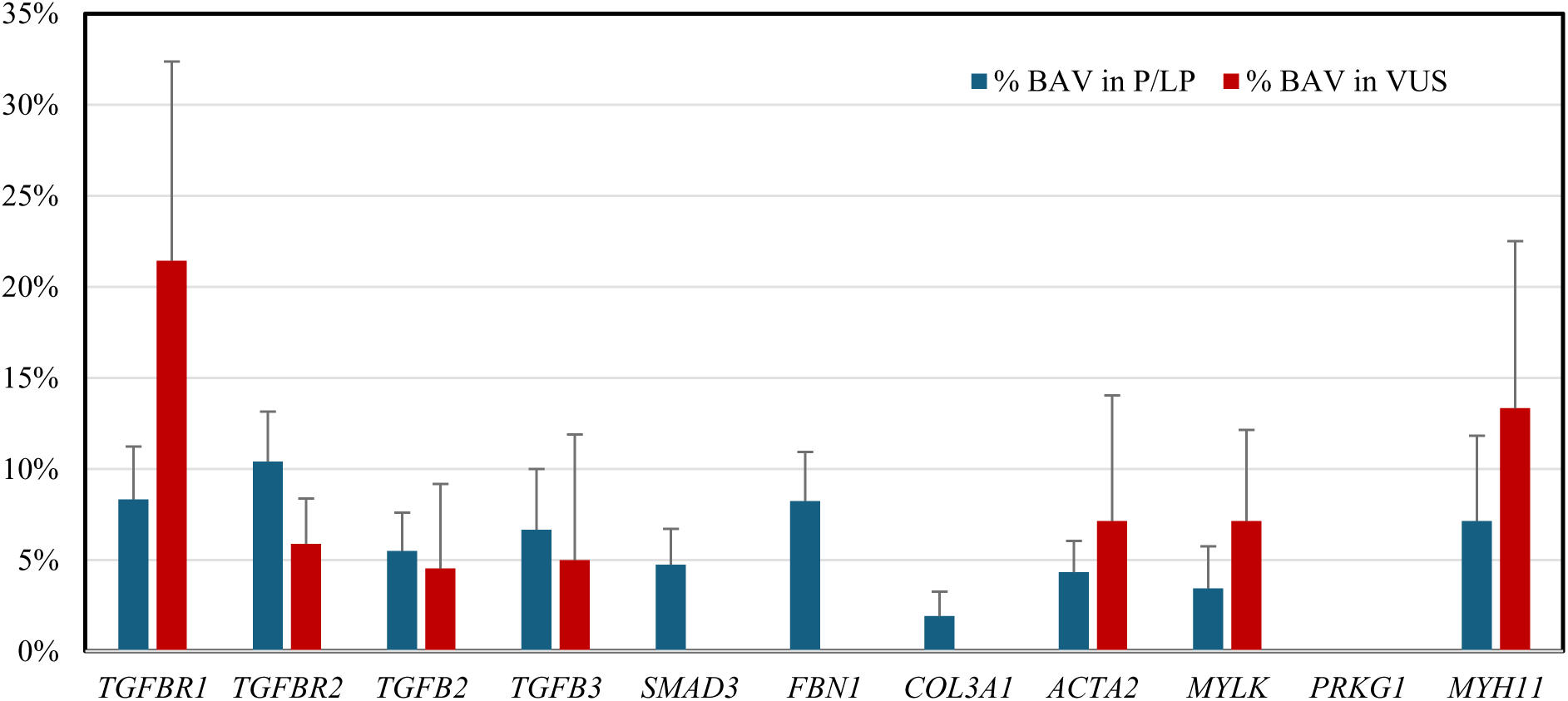
Prevalence of BAV by HTAD variant classification. P/LP: Pathogenic/Likely pathogenic variants. VUS: variants of uncertain significance. *SMAD6, SMAD2, COL5A1, MFAP5, HCN4, SLC2A10, COL5A2, LOX, FLNA, LTBP3, COL12A1, HCN4, SKI, FOXE3, TNXB, COL1A2* PVs were excluded due to fewer than 15 cases total.

**Figure S8:**
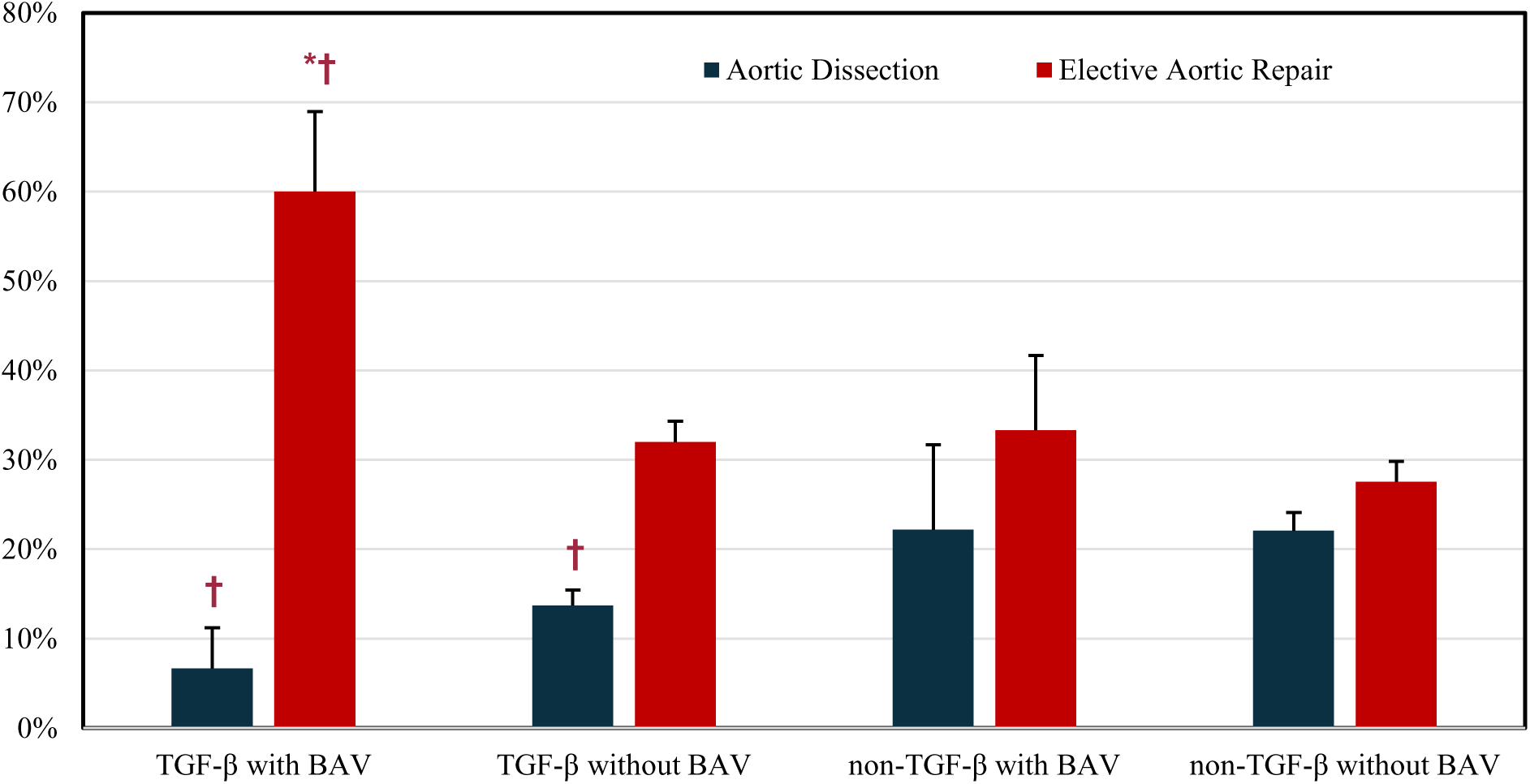
Aortic dissection and elective aortic repair by BAV and TGF-β PV. Asterisk: p<0.05 compared to ‘TGF-β without BAV’ group. Dagger: p<0.05 compared to ‘non-TGF-β without BAV’ group.

